# Evaluation of fractional flow reserve and atherosclerotic plaque characteristics on coronary non-contrast T1-weighted magnetic resonance imaging

**DOI:** 10.1101/2023.07.27.23293293

**Authors:** Hiroki Sugane, Yasuhide Asaumi, Soshiro Ogata, Michito Kimura, Tomoaki Kanaya, Tomoya Hoshi, Akira Sato, Hiroyuki Miura, Yoshiyuki Tomishima, Yoshiaki Morita, Kazuhiro Nakao, Fumiyuku Otsuka, Yu Kataoka, Tomohiro Kawasaki, Kunihiro Nishimura, Jagat Narula, Satoshi Yasuda, Teruo Noguchi

## Abstract

**Background:** The relationship between high-risk coronary plaque characteristics regardless of the severity of lesion stenosis and myocardial ischemia remains unsettled. High-intensity plaques (HIPs) on non-contrast T1-weighted magnetic resonance imaging (T1WI) have been characterized as high-risk coronary plaques. We sought to elucidate whether the presence of coronary HIPs on T1WI influences fractional flow reserve (FFR) in the distal segment of the vessel.

**Methods:** We retrospectively analyzed 232 vessels in 190 patients with chronic stable coronary syndrome who underwent both invasive FFR measurement and coronary T1WI using a multicenter registry. The plaque-to-myocardial signal intensity ratio (PMR) of the most stenotic lesion was evaluated; a coronary plaque with PMR >1.4 was defined as a HIP.

**Results:** The median PMR of coronary plaques on T1WI in vessels with FFR ≤0.80 was significantly higher than that of plaques with FFR >0.80 (1.18 [interquartile range (IQR): 0.96–1.45] vs. 0.97 [IQR: 0.85–1.12]; p<0.001)). Multivariable analysis showed that an increase in PMR of the most stenotic segment is associated with lower FFR (beta-coefficient, –0.051; p<0.001). The presence of coronary HIPs was an independent predictor of FFR ≤0.80 (odds ratio, 5.54; 95% confidence interval, 1.50–20.5; p=0.010).

**Conclusions:** Coronary plaques with high PMR are associated with low FFR in the corresponding vessel, indicating that plaque morphology might influence the degree of myocardial ischemia.

**Trial registration:** UMIN 000029246

**Clinical Perspective:** *What is new?:*

- Incremental of coronary plaque to myocardial signal intensity ratio on T1-weithted magnetic resonance imaging (T1WI), which is represented as instability of coronary plaques was associated with low factional flow reserve (FFR) value in patients with chronic coronary syndrome (CCS).
- The presence of a coronary high-intensity plaque (HIP) detected with T1WI is a strong predictor of low fractional flow reserve (FFR) even after adjusting age, gender, proximal left anterior descending artery, the severity of stenosis in the most stenotic lesion and variables related to plaque volume evaluated on computed tomography angiography.

*What are the clinical implications?:*

- Non-contrast T1WI without contrast media is an anatomy-based, but not ischemia-based, screening method for predicting future coronary events.
- The presence of a coronary HIP on T1WI, which represents a complicated atheroma, is an additional determinant of coronary physiology.

## Introduction

Fractional flow reserve (FFR) is the current reference standard for functional evaluation of myocardial ischemia.^1^ Numerous studies involving coronary computed tomography angiography (CTA) have reported that, in addition to the anatomical severity of coronary lesion stenosis, high-risk plaque features of coronary lesions are associated with low FFR.^2, 3^ Given that a high-risk plaque feature is a potential substrate for future coronary events,^4^ this finding could partly account for the ability of FFR to predict future cardiac events.^5^ High-risk plaques could cause functional stenosis in the presence of hyperemic agents that dilate coronary resistance vessels and possibly uninvolved epicardial coronary artery segments. The lack of vasodilatation at the site of the lesion might result from the Glagovian limit in maximally positively remodeled lesions and endothelial dysfunction.^6, 7^ However, the precise interplay between plaque composition and coronary flow attenuation of lesion stenosis remains to be fully elucidated.

Cardiac magnetic resonance imaging (CMR) of plaques, which is performed without radiation or iodinated contrast exposure, has been used for plaque characterization.^8^ Coronary high-intensity plaques (HIPs) on T1-weighted magnetic resonance imaging (T1WI) are associated with future coronary events in patients with chronic coronary syndrome (CCS).^9–11^ Based on histological studies of coronary or carotid artery atherosclerosis, HIPs on T1WI could represent complicated plaques according to the current AHA classification of atherosclerosis.^8, 12^ However, the exact plaque features responsible for the increased signal intensity on non-contrast T1WI have not yet been fully identified.

We undertook a retrospective analysis of a multicenter registry to investigate the association between invasive FFR and the presence of coronary HIPs with high-risk plaque characteristics in patients with chronic coronary artery disease.

## Methods

### Study population

This study was a retrospective and multicenter observational study (UMIN 000029246). Patients were eligible for this study if they had CCS with planned invasive coronary angiography (ICA) and FFR evaluation for moderately stenotic lesions, i.e., lesions with visually estimated diameter stenosis of 40–80% on angiography due to angina pectoris, presence of myocardial ischemia detected with cardiac radionuclide myocardial perfusion imaging or exercise electrocardiography, coronary stenosis detected on CTA, or history of myocardial infarction. Between August 2012 and November 2019, 1,360 consecutive patients fulfilled these criteria at 4 hospitals: National Cerebral and Cardiovascular Center, Tsukuba University Hospital, Dokkyo Medical University Hospital, and Shin Koga Hospital. Patients were excluded if they had (1) no FFR measurements during the index ICA because of coronary artery stenosis >80% based on visual estimation or obviously thrombotic lesions, (2) FFR measurements for a culprit vessel with a history of previous myocardial infarction, (3) previous coronary stent implantation in a vessel where FFR was measured, (4) stage 4 or 5 chronic kidney disease, (5) devices in the body for which CMR is unsafe, (6) claustrophobia, (7) chronic atrial fibrillation, frequent atrial or ventricular extrasystoles on 12-lead electrocardiography at rest, (8) history of cardiac surgery or thoracic surgery with sternotomy (e.g., coronary artery bypass grafting), (9) hesitancy in undergoing magnetic resonance examination, or (10) poor CMR imaging quality for analysis. The final analysis included 232 vessels in 190 patients who underwent non-contrast T1WI within 6 months before or after ICA with FFR measurement (median, 24 days; interquartile range [IQR], 7– 51) (**Figure 1**). Coronary CTA with both T1WI and FFR values were also available for a sub-analysis of 101 vessels in 80 patients (**Figure S1**). The study protocol was approved by the institutional review board (IRB) or ethics committee of each participating center. Waiver of written informed consent was approved by each IRB or ethics committee because of the retrospective nature of the study. The present study followed the Declaration of Helsinki and the ethical standards of the responsible committee on human experimentation.

**Figure 1.**
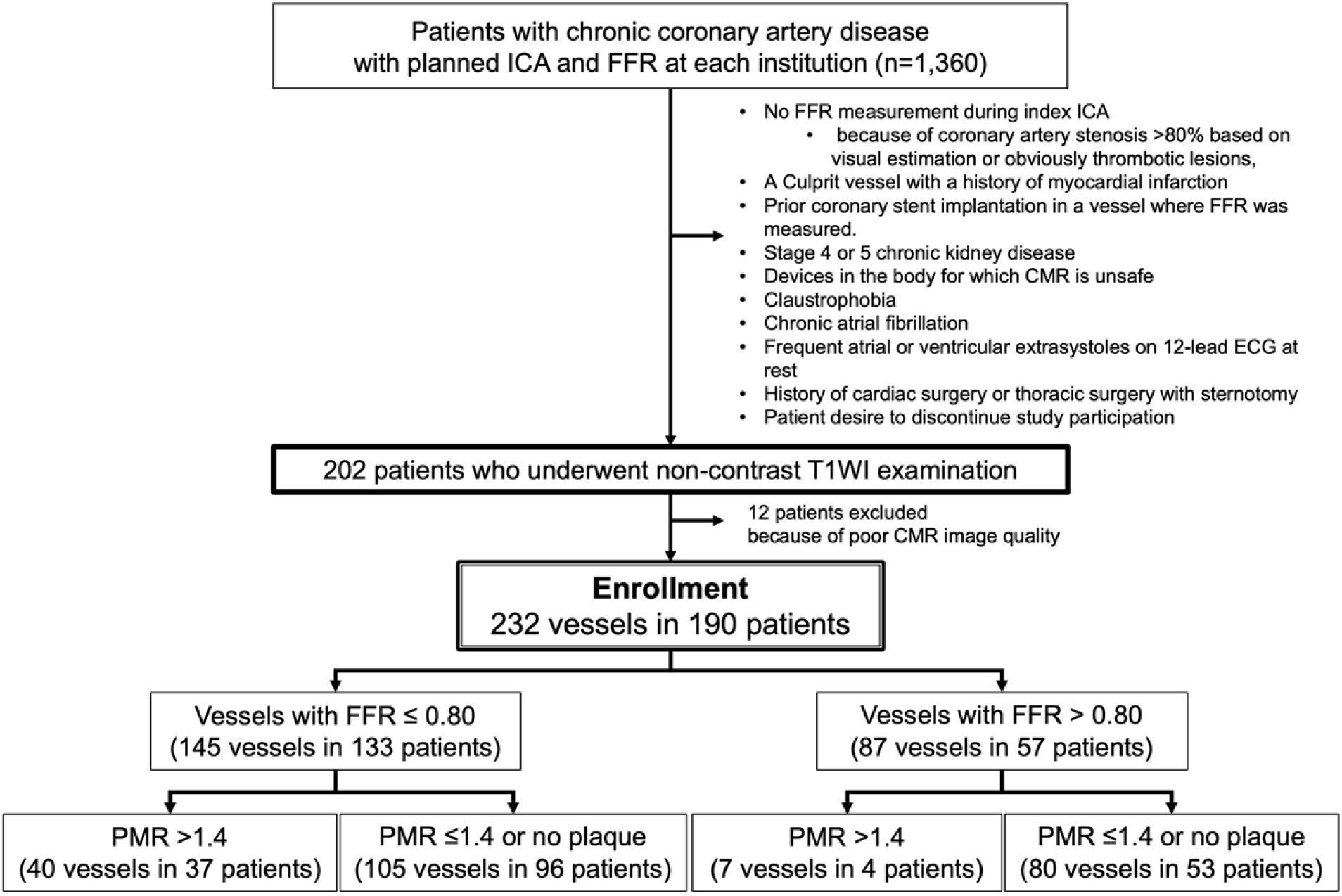
Study flow chart. ICA = invasive coronary angiography, ECG = electrocardiography, FFR = fractional flow reserve, MR = magnetic resonance, T1WI = T1-weighted magnetic resonance imaging.

### ICA and FFR

Selective ICA was performed according to standard practice. Intracoronary nitroglycerin was administered before contrast injection, inducing epicardial vasodilation. The 0.014-inch pressure guide wire (Certus; Abbott, Plymouth, MN) was advanced distal to the stenosis through a 5-Fr or 6-Fr guiding catheter. Equalizing was performed with the guidewire sensor positioned at the guiding catheter tip. FFR was measured during maximal hyperemia induced by an intravenous adenosine infusion administered at 140 μg/kg/min and increased up to 200 μg/kg/min as needed. Low FFR was defined as a FFR value ≤0.80.

### ICA and quantitative coronary angiography

Localization on the coronary tree and quantitative assessment of stenosis severity during ICA was performed offline independently by two expert interventional cardiologists who were blinded to clinical and FFR data. Lesions within the proximal left anterior descending artery (LAD) were defined as being in segment 6, according to the AHA definition of the left coronary artery system.^13^

Quantitative coronary angiography (QCA) was performed using an algorithm that automatically detects arterial contours from acquired ICA digital images (MEDIS, Leiden, the Netherlands). For image calibration, diagnostic or guiding catheters served as the scaling devices to estimate absolute coronary dimensions. The diameter of the catheter was then used to obtain a calibration factor. The automated algorithm was applied to each selected arterial segment. The following absolute coronary dimensions were obtained: the minimal lumen diameter (MLD, smallest diameter of the lumen), reference lumen diameter (RLD, average diameter of the lumen assuming no atherosclerotic disease), and lesion length (distance between the borders separating normal and diseased vessel). Percent diameter stenosis (%DS) was calculated based on these values.^14^

### CMR protocols and CMR image analysis

Non-contrast T1WI was performed on a 3-T magnetic resonance (MR) system with a 32-channel cardiac coil (MAGNETOM Verio; Siemens AG Healthcare Sector, Erlangen, Germany), a 1.5-T MR system (Achieva, Philips Healthcare, Best, The Netherlands) with a 32-element Torso/cardiac phased-array coil, or a 1.5-T MR imager (Intera; Philips Medical Systems, Best, the Netherlands) with 5-element cardiac coils. The procedures for acquiring MR images have been previously described.^9–11, 15^ The plaque-to-myocardium signal intensity ratio (PMR) was defined as the signal intensity of the coronary plaque divided by the signal intensity of the nearby left ventricular myocardium. The methods used to evaluate images of plaques in this study have been described previously.^9–11^ A coronary plaque with PMR >1.4 was defined as a coronary HIP, as reported previously.^9^ The location of the lesion of interest was determined by carefully comparing images using fiduciary points such as side branches. All imaging data were analyzed by two independent cardiologists (YM and TM) who were blinded to clinical data, including FFR.

### Coronary CTA and analysis

Coronary CTA was performed using a dual-source CT scanner (SOMATOM Definition Flash, Siemens Healthcare, Erlangen, Germany). Quantitative lesion analysis was performed using software (Ziostation2, Ziosoft, Tokyo, Japan) that facilitates plaque volume measurement and assessment of the remodeling index,^16^ presence of spotty calcifications, and plaque attenuation in Hounsfield units (HUs). Total plaque volume analysis was performed for the lesion and for the entire vessel of interest. Remodeling index ≥1.1 was interpreted as positive remodeling. The assessment of plaque composition included very low-attenuation non-calcified plaques (VLNCPs) (≤30 HU), low-attenuation non-calcified plaques (LNCPs) (31–50 HU), and non-calcified plaques (NCPs) (51–150 HU). The lumen (151–350 HU) and calcified plaques (CP) (351–1,000 HU) were also defined.^17^ Total non-calcified plaque volume was defined as the sum of VLNCP, LNCP, and NCP plaque volumes. Spotty calcification was defined as calcifications <3 mm in size on focal multiplanar reconstruction images and cross-sectional images.^16, 18^

### Statistical analysis

Continuous variables were presented as medians with interquartile range (IQR); they were compared using the Mann-Whitney U test. Baseline categorical variables were compared using Fisher’s exact test or the chi-square test, as appropriate. Differences in the prevalence of coronary HIPs across four subgroups stratified by FFR value of 0.8 and %DS of 50% on ICA were analyzed using Fisher’s exact test for proportions. Multivariable analyses with a mixed model that included FFR ≤0.80, %DS ≥50%, male gender, and age were conducted to identify factors associated with HIPs in all lesions. Mixed effects models were also used to identify factors associated with FFR values or FFR ≤0.80. A mixed logistic regression model with a random intercept was used to account for the correlation in outcomes between multiple vessels within the same patient. All candidate predictors were first tested for associations in univariable analyses. Subsequently, multivariable stepwise logistic regression with a p value of 0.10 for backward elimination was performed to select the best predictive model using covariates that significantly predicted FFR value or FFR ≤0.80 in the mixed models. Regression coefficients and odds ratios (ORs) with 95% confidence intervals (CIs) were reported as effect sizes for mixed effects models. Intraclass correlation coefficients with 95% CIs were calculated to assess interobserver agreement for PMR on non-contrast T1WI, VLNCP volume, non-calcified plaque volume, and lesion length on CTA. Bland-Altman plots were constructed to visualize agreement. All analyses were performed using Stata 17 SE (Stata Corp LP, College Station, TX) or GraphPad Prism 8 (GraphPad Software, Boston, MA) statistical software. A two-sided p<0.05 was considered statistically significant.

## Results

The clinical characteristics of the study population are summarized in **Table 1**. Among the 190 patients, median age was 70 (IQR, 63–76) years and 77% of patients were male. There were 68 patients with 1 diseased vessel (35.8%), 72 with 2 diseased vessels (37.9%), and 50 with 3 diseased vessels (26.3%). Of those, 149 (78.4%), 40 (21.1%), and 1 (0.5%) underwent 1-, 2-, or 3-vessel FFR evaluation, respectively. In lesion-based analysis, 69% of vessels with FFR measurements were located in the LAD. **Table 2** shows ICA and CMR findings. Median %DS in target plaques on ICA was 49.2% (IQR, 41.3–57.2). FFR in the distal segment of the same vessel was 0.77 (IQR, 0.70–0.83); PMR on T1WI was 1.09 (IQR, 0.90–1.31). Compared with lesions with FFR >0.80, lesions with FFR ≤0.80 were more frequently located in the LAD (83.4% vs. 44.8%, p<0.001), had smaller MLD (1.15 [IQR, 0.96–1.39] vs. 1.46 [IQR, 1.27–1.66] mm; p<0.001), or had longer lesion length (10.8 [IQR, 7.7–14.8] vs. 9.7 [IQR, 5.8–11.3] mm; p=0.022).

**Table 1.** Baseline characteristics of the study patients (n=190)

| Characteristic |  |
| --- | --- |
| Demographic |  |
| Age, years | 70 (63–76) |
| Male | 147 (77.4) |
| Body mass index, kg/m<sup>2</sup> | 23.7 (21.8–26.2) |
| Hypertension | 146 (76.8) |
| Dyslipidemia | 143 (75.3) |
| Diabetes mellitus | 78 (41.1) |
| Current smoking | 24 (12.6) |
| Chronic kidney disease | 42 (22.1) |
| Prior cerebral infarction | 16 (8.4) |
| Peripheral artery disease | 27 (14.2) |
| Prior PCI | 51 (26.9) |
| Prior myocardial infarction | 39 (20.5) |
| Left ventricular ejection fraction, % | 60 (55–63) |
| Number of diseased vessels on ICA (1/2/3) | 68 (35.8) / 72 (37.9) / 50 (26.3) |
| Number of vessels with FFR measurements (1/2/3) | 149 (78.4) / 40 (21.1) / 1 (0.5) |
| Medications |  |
| Aspirin | 163 (85.7) |
| Statin | 151 (80.0) |
| Beta-blocker | 107 (56.3) |
| ACEI or ARB | 81 (42.6) |

Values are n (%) or medians (interquartile range).
ACEI = angiotensin-converting enzyme inhibitor; ARB = angiotensin receptor blocker; FFR = fractional flow reserve; ICA = invasive coronary angiography; PCI = percutaneous coronary intervention.

**Table 2.**
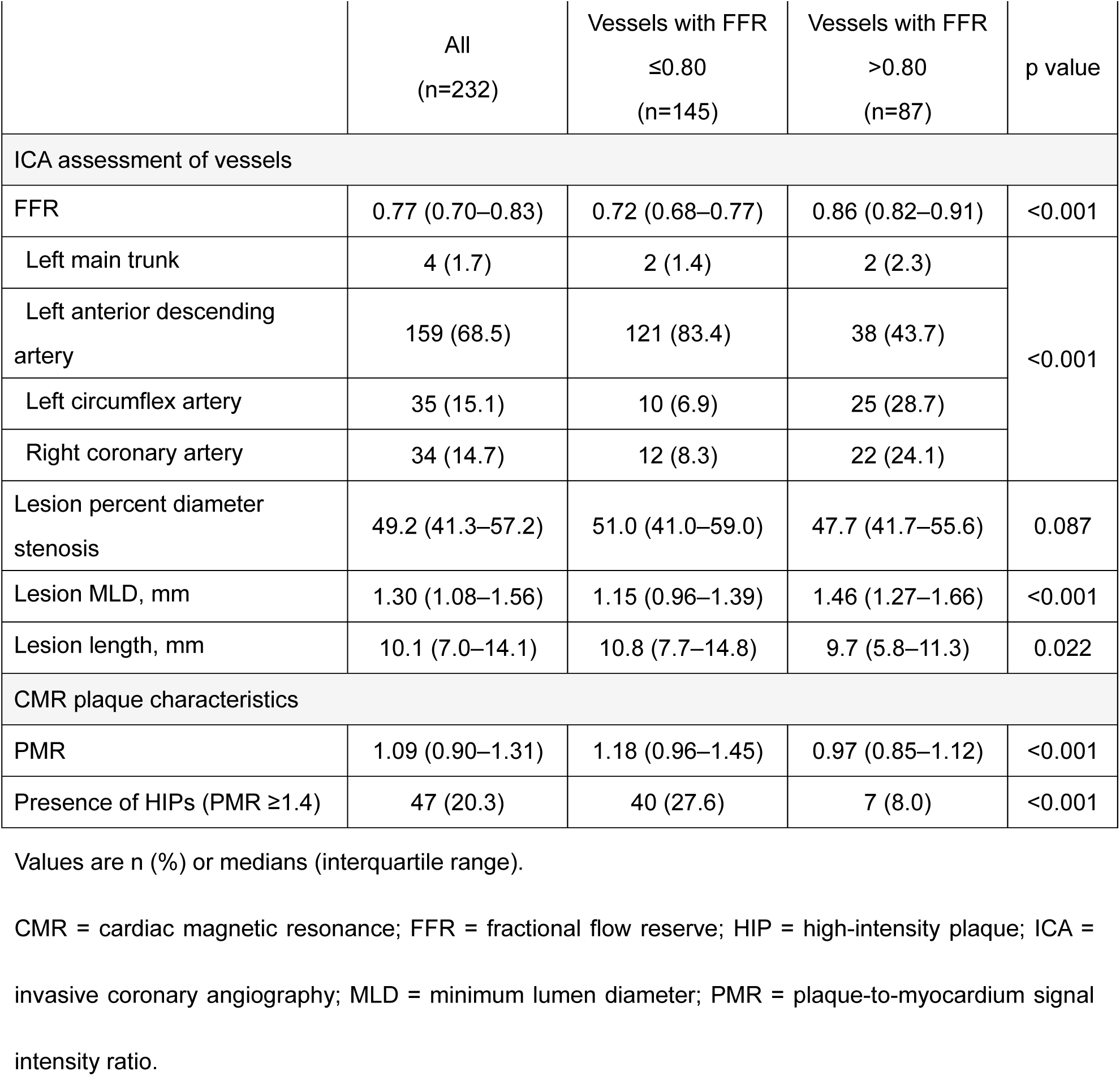
Clinical demographics, ICA findings, and CMR plaque characteristics.

**Figures 2 and 3** show representative cases of coronary non-HIPs and HIPs on T1WI, respectively. In the per-vessel analysis, FFR values in the distal segment of vessels with plaques was inversely correlated with PMR at the lesion (r=-0.275; p<0.001) (**Figure 4A**). PMR at lesions with FFR ≤0.80 was higher than PMR at lesions with FFR >0.8 (1.18 [IQR, 0.96–1.45] vs. 0.97 [IQR, 0.85 –1.12]; p<0.001) (**Figure 4B**). **Figure 4C** is a scatterplot showing the relationships among %DS, FFR, and presence of HIPs in all vessels. There was no correlation between %DS and FFR (r=-0.143). When lesions were subdivided into four groups according to %DS <50% or ≥50% and FFR value ≤0.80 or >0.80, the prevalence of HIPs in the FFR ≤0.80 groups was higher than that in the FFR >0.80 groups, regardless of the degree of coronary stenosis on ICA (p=0.003, Fisher’s exact test: **Figure 4D**). Multivariable logistic regression showed that FFR ≤0.80 is an independent determinant of HIP after adjusting for %DS ≥50%, male gender, and age (OR, 4.21; 95% CI, 1.74–10.2; p=0.001). Multivariable analysis showed proximal LAD lesion (standardized coefficient, –0.046; p<0.001), PMR (standardized coefficient, –0.052; p<0.001), MLD on ICA (standardized coefficient, 0.097; p<0.001), age (standardized coefficient, 0.002; p=0.003), and male gender (standardized coefficient, –0.032; p=0.013) were associated with FFR, but %DS was not (**Table S1**). After dichotomizing FFR based on the cutoff value for myocardial ischemia of 0.80, multivariable analysis showed that proximal LAD lesion (OR, 5.47; 95% CI, 1.53–19.5; p=0.009), PMR (OR, 9.08; 95% CI, 2.54–32.4; p=0.001), and MLD (OR, 0.06; 95% CI, 0.01–0.31, p=0.001) are associated with FFR ≤0.80 even after adjusting for age and male gender (**Table 3, Model I**). PMR >1.4 was also a predictor of FFR ≤0.80 (OR, 5.54; 95% CI, 1.50–20.5, p=0.010) (**Table 3, Model II**).

**Figure 2.**
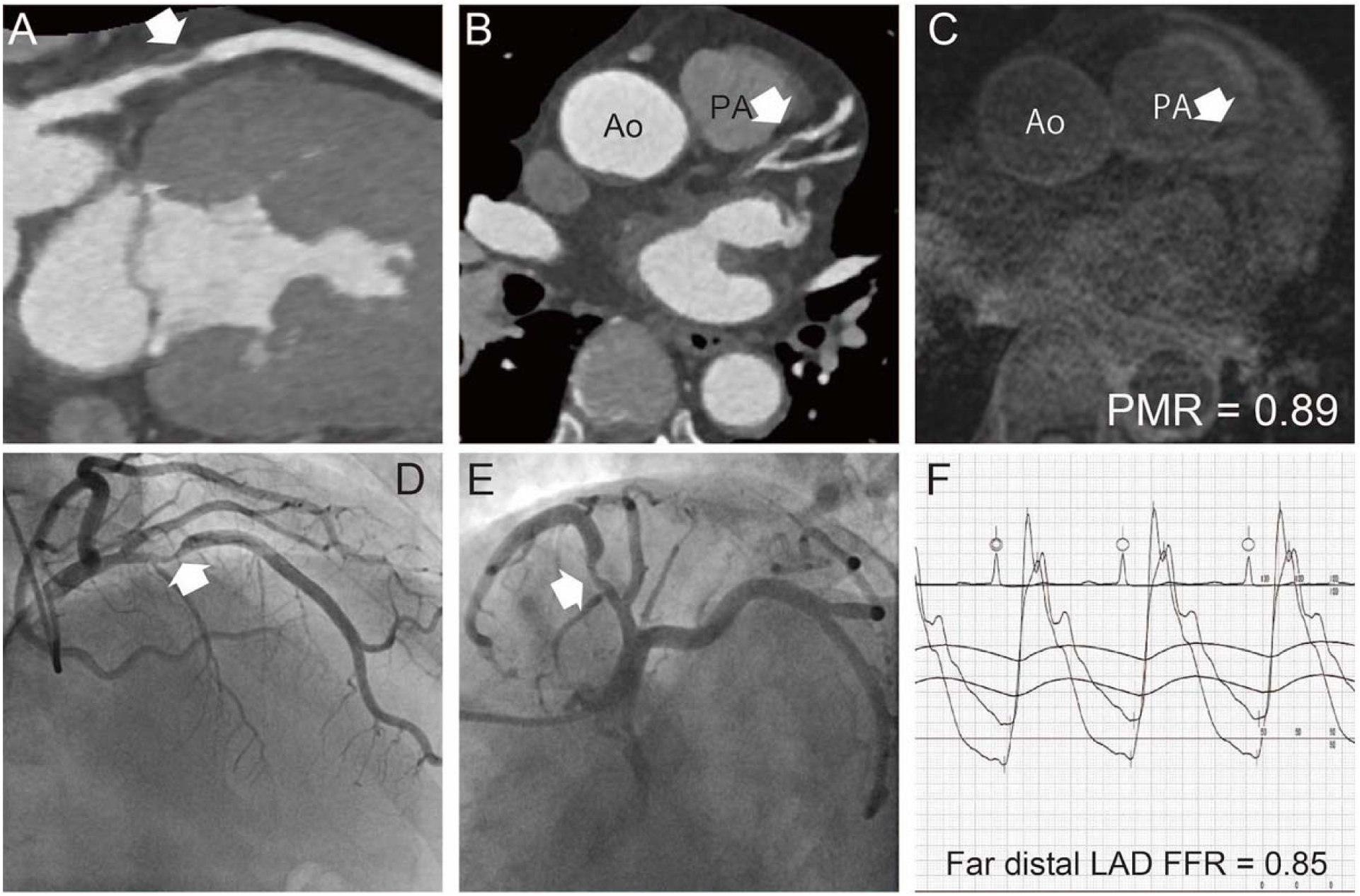
Representative FFR and non-HIP lesion on T1WI, CTA, coronary angiography. Coronary CTA showed an intermediate stenosis in a proximal lesion in the LAD (A and B, white arrows). The PMR of this lesion on non-contrast T1WI was 0.89 (white arrow, C). Invasive coronary angiography demonstrated intermediate stenosis in a proximal lesion in the LAD (D and E, white arrows). FFR in the vessel distal to the corresponding plaque was 0.85 (F). Ao = ascending aorta, CTA = computed tomography angiography, FFR = fractional flow reserve, HIP = high-intensity plaque, LAD = left anterior descending artery, PA = pulmonary artery, PMR = plaque-to-myocardial signal intensity ratio, T1WI = T1-weighted magnetic resonance imaging.

**Figure 3.**
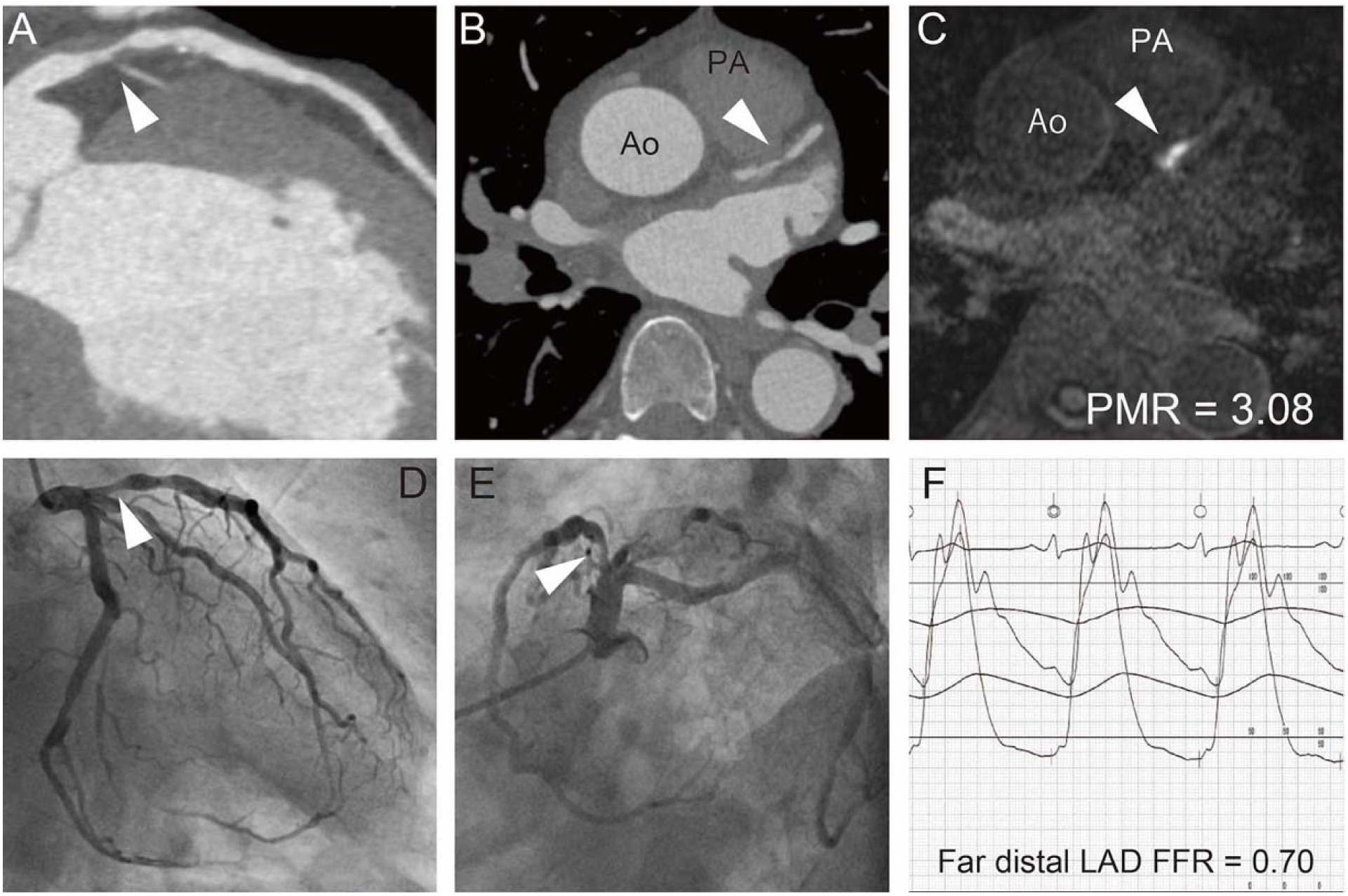
Representative FFR and HIP on T1WI, CTA, and coronary angiography. Coronary computed tomography angiography showed moderate stenosis in an ostial lesion in the LAD (A and B, white arrowheads). The PMR value of this lesion was 3.08 (white arrowhead, C). Invasive coronary angiography demonstrated moderate stenosis in an ostial lesion in the LAD (D and E, white arrowheads). The FFR value in a vessel distal to the corresponding plaque was 0.70 (F). Ao = ascending aorta, CTA = computed tomography angiography, FFR = fractional flow reserve, HIP = high-intensity plaque, LAD = left anterior descending artery, PA = pulmonary artery, PMR = plaque-to-myocardial signal intensity ratio, T1WI = T1-weighted magnetic resonance imaging.

**Figure 4.**
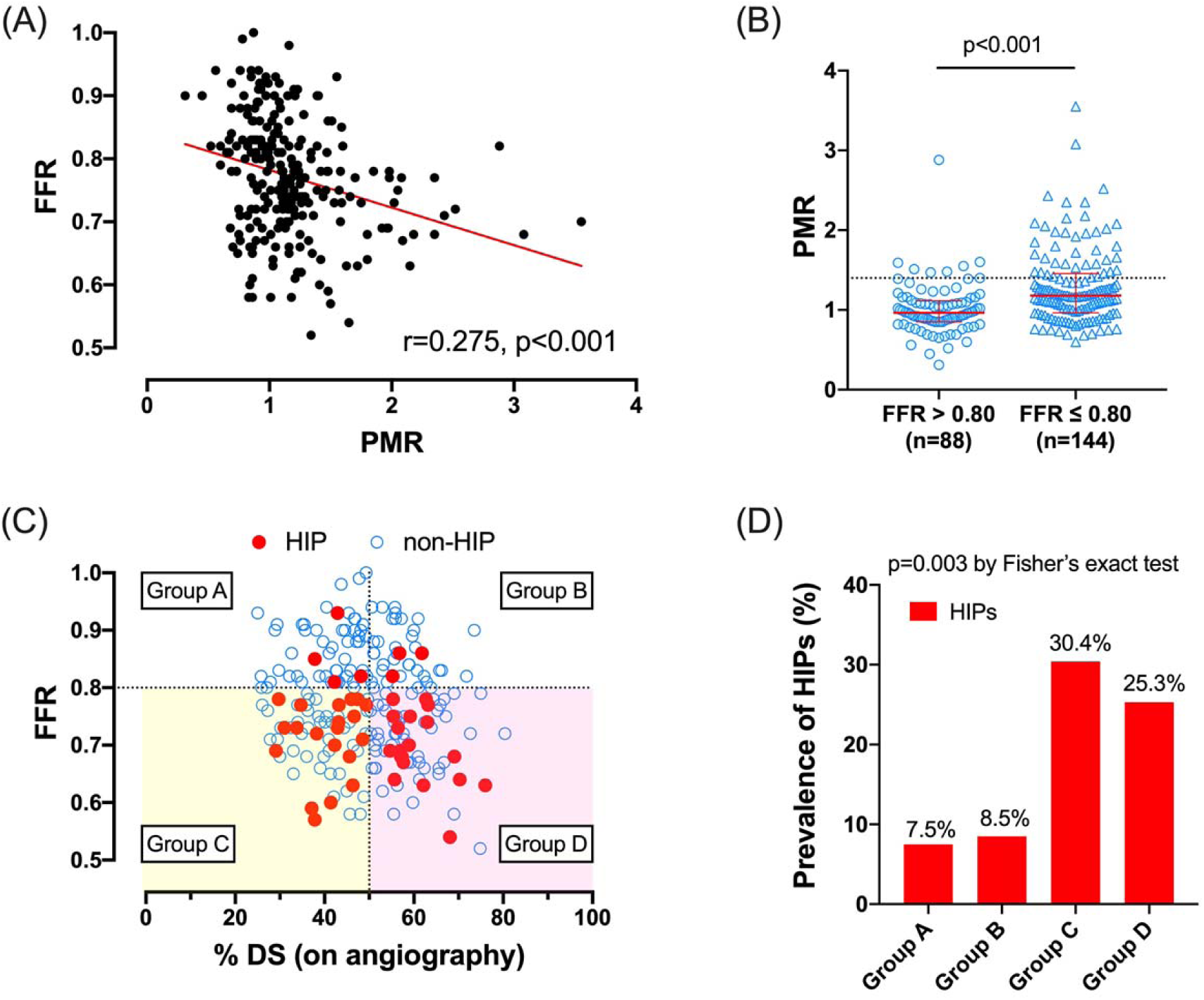
Correlation between PMR and FFR or %DS among all lesions. (**A**) Scatter plots demonstrating the relationship between PMR and FFR **(B)** Differences in PMR between lesions with FFR ≤0.80 and lesions with FFR >0.80 **(C)** Scatter plots demonstrating the relationships among FFR, %DS, and PMR >1.4 **(D)** Prevalence of HIPs in groups with FFR >0.80 and %DS ≤50% (Group A), FFR >0.80 and %DS >50% (Group B), FFR ≤0.80 and %DS ≤50% (Group C), or FFR ≤0.80 and %DS >50% (Group D) FFR = fractional flow reserve, HIP = high-intensity plaque, PMR = plaque-to-myocardial signal intensity ratio, %DS = percent diameter stenosis.

**Table 3.**
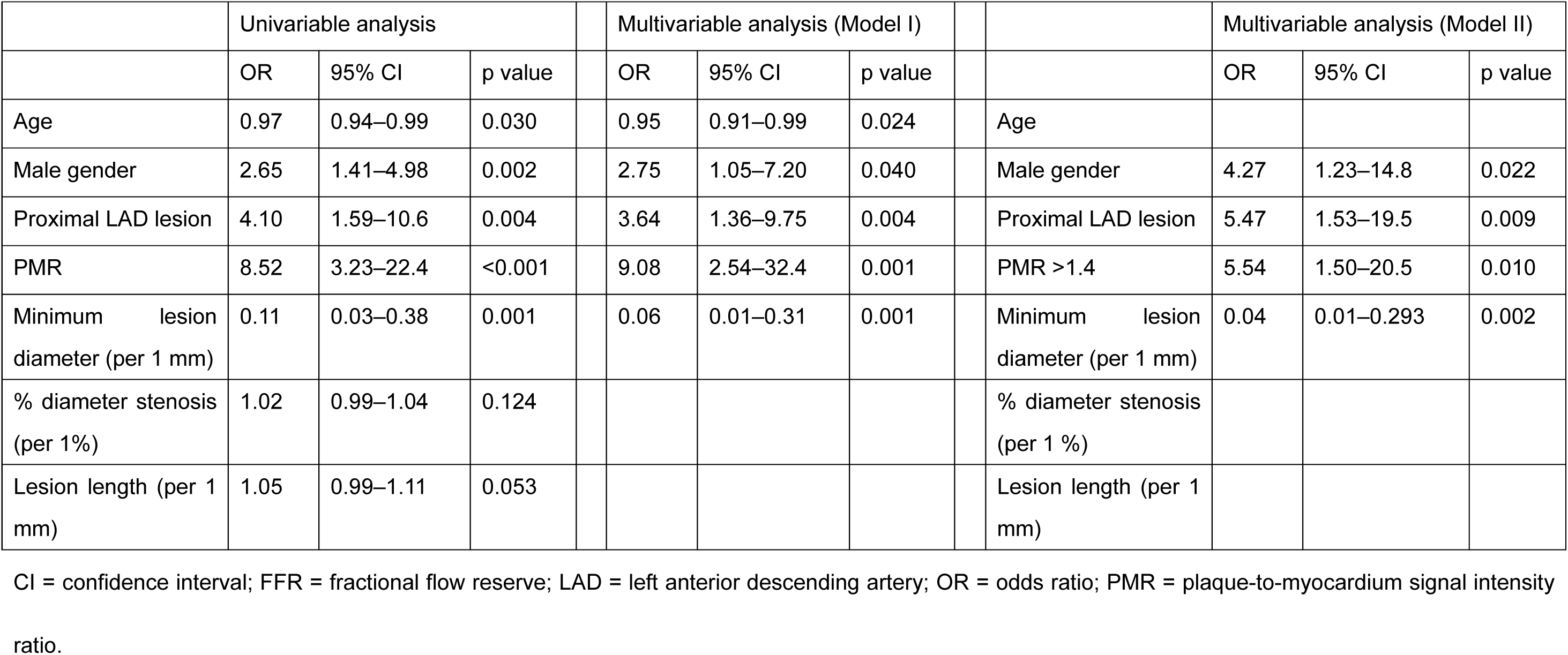
Determinants of FFR ≤0.8 (232 lesions from 190 patients)

To clarify the effects of plaque volume on FFR beyond the effect of coronary stenosis, we explored the relationships among plaque volume from CTA, FFR, and coronary HIP in 101 vessels from 80 patients (**Figure S1**). In the per-vessel analysis (**Table 4**), the prevalence of coronary HIPs on T1W and LAD lesions were higher in vessels with FFR ≤0.80. Vessels with FFR ≤0.80 also tended to have a higher VLNCP volume than vessels with FFR ≤0.80, regardless of the degree of coronary stenosis. Multivariable analysis showed that FFR is inversely correlated with PMR (beta-coefficient, –0.031; 95% CI, –0.057 to –0.006; p=0.017) and proximal LAD lesion (beta-coefficient, –0.055; 95% CI, –0.082 to –0.027, p<0.001) after adjusting for age, male gender, and variables related to plaque volume and CTA-verified extent of coronary stenosis (**Table S2**). Even after stratification based on FFR of 0.80 for the presence of myocardial ischemia and adjusting for age, male gender, proximal LAD lesion, and CTA-derived variables, higher PMR value or PMR >1.4 were independently associated with FFR ≤0.80 (OR, 4.06; 95% CI, 1.17–14.1; p=0.028, and OR, 5.75; 95% CI, 1.33–24.8; p=0.019, respectively) (**Table 5**).

**Table 4.** Clinical demographics, ICA findings, CMR plaque characteristics, and CTA findings (Study 2: CTA sub-study)

|  | Vessels with FFR<br>≤0.80 (n=58) | Vessels with FFR<br>>0.80 (n=43) | p<br>value |
| --- | --- | --- | --- |
| ICA assessment of vessels |  |  |  |
| FFR | 0.73 (0.69–0.77) | 0.86 (0.82–0.91) | <0.001 |
| Left main trunk | 1 (1.7) | 2 (4.7) | <0.001 |
| Left anterior descending artery | 46 (79.3) | 17 (41.7) |  |
| Left circumflex artery | 5 (8.6) | 10 (23.3) |  |
| Right coronary artery | 6 (10.3) | 13 (30.2) |  |
| Lesion percent diameter stenosis, % | 51 (39.6–59.7) | 49.5 (43.2–56.8) | 0.773 |
| Lesion MLD, mm | 1.3 (1.1–1.5) | 1.4 (1.2–1.7) | 0.069 |
| Lesion length, mm | 10.1 (7.0–15.0) | 10.9 (6.9–13.4) | 0.642 |
| CMR plaque characteristics |  |  |  |
| PMR | 1.17 (0.95–1.54) | 0.95 (0.86–1.08) | <0.001 |
| Presence of HIPs (PMR >1.4) | 17 (29) | 3 (7) | 0.003 |
| CTA findings |  |  |  |
| Stenosis, % | 52.2 (42.7–59.7) | 48.5 (42.9–56.9) | 0.559 |
| Plaque length, mm | 20.7 (16.0–24.3) | 18.0 (12.0–23.0) | 0.065 |
| Minimal lesion diameter, mm | 1.5 (1.1–1.6) | 1.5 (1.3–1.8) |  |
| Very low-attenuation non-calcified plaque volume, mm<sup>3</sup> | 25.0 (15.8–39.5) | 18.0 (11.0–26.0) | 0.066 |
| Non-calcified plaque volume, mm<sup>3</sup> | 127 (82.5–167) | 99 (62–147) | 0.081 |
| Calcified plaque, volume, mm<sup>3</sup> | 10.8 (1.0–33.0) | 11.0 (2.0–39.0) | 0.565 |
| Lumen area, mm<sup>3</sup> | 123 (81–172) | 119 (80–188) | 0.885 |
| Plaque burden, % | 54.1 (47.7–60.0) | 50.6 (42.7–56.9) | 0.091 |
| Remodeling index | 1.12 (0.98–1.33) | 1.05 (0.95–1.20) | 0.322 |
| Positive remodeling | 30 (51) | 20 (47) | 0.604 |
| Spotty calcification | 20 (34) | 14 (33) | 0.839 |

Values are n (%) or medians (interquartile range).
CTA = computed tomography angiography; CMR = cardiac magnetic resonance; FFR= fractional flow reserve; HIP = high-intensity plaque; ICA = invasive coronary angiography; MLD = minimum lumen diameter; PMR = plaque-to-myocardium signal intensity ratio.

**Table 5.** Determinants of FFR ≤0.80 (101 lesions in 80 patients)

|  | Univariable analysis |  |  |  | Multivariable analysis: Model I |  |  |  | Model II |  |  |
| --- | --- | --- | --- | --- | --- | --- | --- | --- | --- | --- | --- |
|  | OR | 95% CI | p value |  | OR | 95% CI | p value |  | OR | 95% CI | p value |
| Age | 0.89 | 0.84–0.95 | <0.001 |  | 0.88 | 0.83–0.94 | <0.001 | Age | 0.88 | 0.83–0.94 | <0.001 |
| Male gender | 2.39 | 1.36–21.14 | 0.017 |  |  |  |  | Male gender |  |  |  |
| Proximal LAD lesion | 1.98 | 1.02–25.0 | 0.048 |  | 4.42 | 1.63–12.0 | 0.003 | Proximal LAD lesion | 4.49 | 1.66–12.1 | 0.003 |
| PMR | 4.36 | 1.30–14.62 | 0.017 |  | 4.06 | 1.17–14.1 | 0.028 | PMR >1.4 | 5.75 | 1.33–24.8 | 0.019 |
| Very low-attenuation non-calcified plaque volume | 1.01 | 0.99–1.04 | 0.325 |  |  |  |  | Very low-attenuation non-calcified plaque volume |  |  |  |
| Total non-calcified plaque volume | 1.00 | 0.99–1.01 | 0.254 |  |  |  |  | Total non-calcified plaque volume |  |  |  |
| Positive remodeling | 1.20 | 0.51–2.84 | 0.671 |  |  |  |  | Positive remodeling |  |  |  |
| Spotty calcification | 1.09 | 0.43–2.75 | 0.857 |  |  |  |  | Spotty calcification |  |  |  |
| MLD on CTA | 0.28 | 0.07–1.16 | 0.079 |  |  |  |  | MLD on CTA |  |  |  |
| % diameter stenosis on CTA | 1.01 | 0.98–1.05 | 0.541 |  |  |  |  | % diameter stenosis on CTA |  |  |  |
| Lesion length on CTA | 1.06 | 1.00–1.12 | 0.067 |  |  |  |  | Lesion length on CTA |  |  |  |

CI = confidence interval; CTA = computed tomography angiography; FFR = fractional flow reserve; MLD = minimum lumen diameter; OR = odds ratio;
PMR = plaque-to-myocardium signal intensity ratio.

## Reproducibility

A total of 42 lesions from 42 patients who underwent T1WI and 12 lesions from 9 patients who underwent CTA were reanalyzed to test interobserver variability. For PMR, interobserver reproducibility, expressed as an ICC, was 0.950 (95% CI, 0.917–0.970). For VLNCP volume, total NCP volume, and lesion length on CTA, ICCs were 0.993 (95% CI, 0.976–0.998), 0.923 (95% CI, 0.632–0.979), and 0.938 (95% CI, 0.812–0.981), respectively. The Bland-Altman plots showed small differences in variables measured by two observers (**Figure S2**).

## Discussion

This study demonstrated an association between high-risk coronary plaque characteristics detected on non-contrast T1WI and myocardial ischemia in corresponding vessels evaluated with invasive FFR measurements, regardless of the extent of coronary diameter stenosis in patients with chronic coronary artery disease.

Although several studies have previously shown a similar relationship between plaque characteristics and coronary flow attenuation regardless of the degree of coronary stenosis, most of these studies were derived from CTA analyses. Bauer et al. showed that a higher burden from low attenuated non-calcified plaques is associated with the presence of myocardial ischemia across from a corresponding coronary atheroma.^19^ Park et al. demonstrated that the presence of a coronary atheroma with lower attenuation, positive vessel remodeling, or both is associated with the presence of myocardial ischemia.^2^ Larger volume of lower-attenuation plaques with positive vessel remodeling on CTA is associated with future coronary events in patients with suspected coronary artery disease.^4, 16^ Taken together, coronary plaques with high-risk features might therefore be susceptible to hemodynamic perturbances in coronary flow.

In the present study, coronary HIPs with high PMR were associated with low FFR in the distal segment of corresponding vessels. As shown in **Figure 4**, coronary PMR was inversely correlated with FFR and the presence of coronary HIPs with high PMR was associated with FFR ≤0.80, regardless of %DS and lesion length on ICA. Moreover, the prevalence of coronary HIPs was significantly higher in segments with FFR ≤0.80 than in those with FFR >0.80, regardless of stenosis severity (**Figure 4C-D** and **Table 2**). To predict future coronary events in patients with CCS as part of risk stratification, both ischemia-based approaches (e.g., flow disturbance from coronary stenosis) and plaque characteristics-based approaches (e.g., plaque characteristics) are currently used. According to the Providing Regional Observations to Study Predictors of Events in Coronary Tree (PROSPECT) study, both coronary stenosis and plaque characteristics play important roles in the development of future coronary events.^20^ FFR measurement is the standard reference for evaluating vessel-based myocardial ischemia during ICA.^21, 22^ Low FFR is associated with coronary events.^23^ During the development of coronary atherosclerosis and coronary events, coronary flow disturbance and plaque characteristics with high-risk features overlap with each other. Taken together, non-contrast T1WI without contrast media is an anatomy-based, but not ischemia-based, screening method for predicting future coronary events.^9^

This study also showed that proximal LAD lesion MLD of the lesion are associated with the functional severity of the stenosis, but not %DS. Previous studies involving more patients that examined the relationship between %DS and FFR showed that FFR is moderately correlated with %DS.^24^ However, one-third of patients had discordance between %DS of 50% and FFR of 0.80,^24, 25^ which suggests FFR ≤0.80 and %DS on ICA might result in low predictive accuracy. In particular, a higher prevalence of positive mismatch (FFR ≤0.80 in lesions with %DS <50%) and lower prevalence of negative mismatch (FFR >0.80 in lesions with %DS ≥50%) were observed in LAD lesions versus non-LAD lesions.^25, 26^ A large territory of myocardium is supplied by the LAD ^27^, which leads to a greater likelihood of functional significance in LAD lesions than in non-LAD lesions.^24, 26, 28^

The mechanism underlying the association between coronary HIPs and myocardial ischemia remains unknown. As shown in **Table 5** and **Table S2**, we examined independent determinants of low FFR, including both T1WI and CTA parameters to adjust for plaque volume. From this analysis, an incremental increase in coronary PMR on non-contrast T1WI was an independent determinant of FFR, even after the volume of low-attenuation plaques in the corresponding segment on CTA was taken into account. Lesions with coronary HIPs have larger CTA-verified necrotic cores and positive vessel remodeling.^15, 18^ Regarding luminal geometry in atherosclerotic lesions, positive remodeling with a large necrotic core at the site of the lesion represents extraluminal expansion and a stretched smooth muscle layer, which might restrict further vessel dilation according to the limits of the Glagovian phenomenon in hyperemia.^29^ The presence of plaque rupture, which is frequently observed in high-risk plaques,^30^ might contribute to turbulent flow at the lesion and reduce myocardial blood flow independent of the degree of stenosis in the distal segment of the lesion.^26, 31^ This mechanism is also supported by more severe endothelial dysfunction in target vessels harboring a high total plaque and necrotic core volume.^7^ Based on studies that evaluated associations between histological findings in carotid and coronary atherosclerosis and non-contrast T1WI findings, lesions with coronary HIPs might be associated with complicated atherosclerotic plaque.^8, 12, 32^ Our present findings suggest that the presence of a HIP, which represents a complicated atheroma, is an additional determinant of coronary physiology. However, because of the limited spatial resolution or parametric mapping capability of T1WI to identify individual plaque components, the plaque component reflected by PMR as evaluated by non-contrast T1WI has not been adequately identified.

This study has several limitations. First, although our study was a multicenter study, the number of enrolled vessels or patients was small; the number of lesions with HIPs was also relatively small (n=46; 19.8%). Second, FFR measurements for LAD lesions comprised 68.5% (159/232 vessels) of all FFR measurements and vessels with FFR ≤0.80 accounted for 62.5% (145/232 vessels) of vessels in the present study. FFR measurements are dependent on operators’ visual assessment to identify intermediate stenosis with FFR indication. In addition, the operators’ knowledge that LAD lesions are more susceptible to lower FFR than non-LAD lesions with the same degree of intermediate stenosis might have led to a higher rate of FFR measurement in LAD lesions than in non-LAD lesions. These facts might have resulted in selection bias.

In conclusion, coronary plaques with high PMR are associated with low FFR, indicating that plaque morphology might influence the degree of myocardial ischemia.

## Declarations

### Ethics approval and consent to participate

This study protocol was approved by the institutional review board of National Cerebral and Cardiovascular Center (M28-141) and the institutional review board or ethics committee of each participating center. Waiver of written informed consent was approved by each institutional review board or ethics committee because of the retrospective nature of the study.

## Consent for publication

Not applicable. Only non-identifiable data was used.

## Availability of data and materials

The datasets used and/or analyzed during the current study are available from the corresponding author on reasonable request.

## Authors’ contributions

YA and TN conceived and designed this study. HS, YA, TK, and TN implemented the study protocol, performed data analysis, and wrote the manuscript. TK, TH, HM, and YM set up the CMR sequence and performed CMR image analysis. AS, KN, TH, AS, and TK supervised CMR image analysis. HM and YT performed CTA and ICA analyses. SO and KN performed statistical analysis. FO, YK, TK, JN, and SY critically revised the manuscript. All authors read and approved the final manuscript.

## Abbreviations

AHA: the American Heart Association
CCS: chronic coronary syndrome
CMR: cardiac magnetic resonance imaging
CTA: computed tomography angiography
FFR: fractional flow reserve
HIP: high-intensity plaque
ICA: invasive coronary angiography
LAD: left anterior descending artery
MR: magnetic resonance
PMR: plaque-to-myocardial signal intensity ratio
QCA: quantitative coronary angiography
T1WI: T1-weighted magnetic resonance imaging

## Acknowledgements

None

